# Expected reduction in obesity of a 20% and 30% tax to Sugar-Sweetened-Beverages in Brazil: a modeling study

**DOI:** 10.1101/2023.01.25.23285038

**Authors:** Ana Basto-Abreu, Rossana Torres-Alvarez, Tonatiuh Barrientos-Gutierrez, Paula Pereda, Ana Clara Duran

## Abstract

**Background:** The consumption of sugar-sweetened beverages (SSB) is associated with obesity, metabolic diseases, and incremental healthcare costs. Given their health consequences, the World Health Organization (WHO) recommended that countries implement taxes to SSB. Over the last 10 years, Brazil has almost doubled its obesity prevalence, yet, in 2016, the Brazilian government cut 23 percentage points to existing federal SSB taxes to their current 4%. To simulate the potential impact of updating the fiscal policy towards SSB in Brazil, we aimed to estimate the price-elasticity of SSB and model the potential impact of a new 20 or 30% excise SSB tax on consumption, obesity prevalence, and healthcare costs.

**Methods and Findings:** Using household purchases data from the Brazilian Household Budget Survey (POF) from 2017/2018, we estimated constant elasticity regressions using a log-log specification by income level for all beverage categories: (1) sugar-sweetened beverages, (2) alcoholic beverages, (3) other beverages, and (4) low-calorie sweetened beverages. We estimated baseline intake for each beverage group using 24h dietary recall data from POF 2017/2018. Applying the price and cross-price elasticities to the baseline intake, we obtained changes in caloric intake. The caloric reduction was introduced into an individual dynamic model to estimate changes in weight and obesity prevalence. By multiplying the reduction in obesity cases during 10 years by the obesity costs per capita, we predicted the reduction in obesity costs attributable to the sweetened beverage tax. SSB price elasticities were higher in the low (−1.24) than in the high-income tertile (−1.13); cross-price elasticities suggest SSB were weakly substituted by milk, water, and 100% fruit juices. We estimated a caloric change of -17.3 kcal/day/person under a 20% excise tax and -25.9 kcal/day/person for a 30% tax. 10 years after implementation, a 20% tax is expected to reduce obesity prevalence by 6.7% and 9.1% for a 30% tax. These reductions translate into a -2.8 million and -3.8 million obesity cases for a 20 and 30% tax, respectively, and a reduction of $US -257.4 million and $US -345.9 million obesity costs over 10 years for a 20 and 30% tax, respectively.

**Conclusions:** Adding a 20 to 30% excise tax on top of Brazil’s current federal tax in Brazil could help reduce the consumption of ultra-processed beverages, empty calories, and body weight while avoiding large health-related costs. Given the recent cuts to SSB taxes in Brazil, a program to revise and implement excise taxes could prove beneficial for the Brazilian population.

## INTRODUCTION

Consumption of sugar-sweetened beverages (SSB) is associated with increased caloric intake, weight gain, and the development of multiple chronic diseases, such as diabetes, metabolic syndrome, and cancer.[1–3] Governments worldwide are establishing structural interventions to improve consumers’ choices. Following the experience of alcohol and tobacco, taxes to SSB have gained international attention as a potentially effective measure to reduce consumption and improve health.[4] A recent systematic review analyzed the impact of SSB taxes in six jurisdictions, finding on average that a 10% tax on SSBs produces a 10% decrease in consumption.[5] Recently, the World Health Organization recommended that countries implement a 20% tax on sugary beverages to improve diet and reduce chronic diseases.[6]

Latin America has been at the forefront of SSB taxes.[7] In 2014, Mexico implemented a 10% SSB tax that reduced consumption by 9.7% two years after implementation.[8] Using a dynamic weight change model, we estimated that the 10% SSB tax in Mexico would translate into a 2.54% reduction in the prevalence of obesity in adults over ten years.[9,10]. Further simulation studies estimated important reductions in cardiovascular disease,[11] child body weight,[10,11] and obesity-related cancers.[12] In 2016, Chile implemented an integral package of interventions to reduce SSB consumption, including increasing an existing 13% tax to 18%,[13] front-of-package warning labels, restricted child-directed marketing, and banning sales of SSB and unhealthy foods in schools.[14] By 2017, regulated beverage consumption decreased 23.4%, while caloric content decreased 27.5%. SSB taxes and regulations in the Latin American region have proven to reduce consumption and are expected to produce important health benefits.

In 2010, Brazil’s SSB per capita consumption was estimated at 142 ml/day/person.[15] Between 2009 and 2019, the prevalence of obesity in the country increased from 11.8% to 20.3% across all ages, but particularly among young adults,[16] where SSB consumption is more frequent.[15] Despite having high consumption levels, Brazil reduced SSB federal taxes in 2016 by 23 percentage points, and the current SSB federal tax in the country is 4%.[17] Recently, the government and academics have discussed the need to increase SSB taxes to at least 20%.[6,17]

We aimed to estimate the potential impact of an SSB tax in Brazil as a tax aiming at disincentivizing sugar-sweetened industrialized beverages. First, we estimated own- and cross-price elasticities for industrialized SSB and other beverages assuming new 20 and 30% excise taxes were added to the existing federal tax. Then, we used a dynamic simulation model to estimate the expected impact of the taxes on body weight in the adult Brazilian population. Prevented obesity cases were then linked to obesity costs so we could estimate the direct healthcare costs averted by such intervention over a 10-year period.

## METHODS

### Model overview

We used a simulation model to estimate the potential impact of a sweetened beverages tax on obesity and obesity costs in Brazil (Figure 1) as the country is discussing its implementation. First, we used purchases data to estimate own- and cross-price elasticities for industrialized SSB in Brazil; then, we assumed that the expected reduction in purchases is translated into a proportional reduction in consumed calories. Using a dynamic weight change model, we estimated changes in body mass index (BMI) and obesity prevalence over 10 years, then linked to per capita obesity costs to estimate the expected direct costs averted. We simulated two tax scenarios – using a 20 and a 30% SSB tax –, and we considered changes in SSB intake only and the potential caloric substitution by other beverages based on the cross-price elasticities.

**Fig 1.**
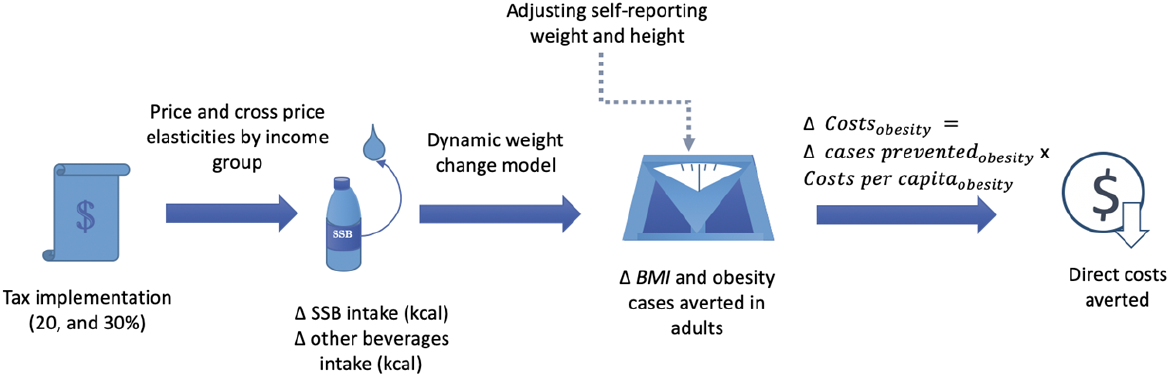
Representation of the simulation strategy. Data sources: POF 2017/2018, PNS 2019, Hall et al.2011, Nilson, 2020, Ward. 2019. Brazilian Institute of Geography and Statistics (IBGE).

### Price elasticities by income level

To estimate the price elasticities of SSB by income level, we used the Brazilian Household Budget Survey (*Pesquisa de Orçamentos Familiares - POF*) from 2017-2018 (POF 2017- 2018) collected by the Brazilian Institute of Geography and Statistics (IBGE). POF 2017-2018 is representative of Brazil, its states, and metropolitan regions. Within POF, we used the purchase database, which registers all food purchases of a sample of 57,920 Brazilian households within a week (representing 59,783,430 Brazilian households). We considered POF survey sampling weights in all estimates.

POF data include food acquisition - in total quantity (in kg) - and monetary value (in Brazilian reais). We categorized the beverage items of POF by using the NOVA classification into the following categories: (1) sugar-sweetened beverages, (2) alcoholic beverages, (3) other beverages, and (4) low-calorie sweetened beverages. Table A in S1 Appendix describes the beverage items included in each category.

We also categorized food items to include food prices as controls in the demand regressions. Other food groups included: (i) fruits, (ii) *in natura* food, (iii) *in natura* meat, (iv) snacks, (v) sugar, (vi) other processed food, and (vii) other food items. Prices were calculated as unit values, calculated as total expenditure divided by total quantity consumed (implicit price). If a household reported zero consumption of an item, we approximated its price by the median of the neighboring regions. We estimated basic constant elasticity regressions using a log-log specification for all beverage categories:

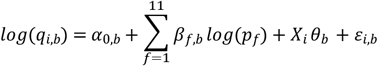

In which *q*_*i,b*_ is the total quantity (in kg) of beverage *b* acquired by household *i*. We included the price logarithm for all 11 categories of food and beverages (indexed by *f*). We also included several sociodemographic variables as control variables (matrix *X*), such as (i) variables of the household head (gender, years of schooling, race, and marital status), and variables of the household (total members of the household, total children within the household, and logarithm of the income in levels and quadratic).

We considered different price elasticities by income level. Income levels were categorized using the tertiles (*T*) of the distribution of income per capita of the households (*Tertlncome*_*T*_) calculated using sampling weights. Thus, the basic regression was modified according to the following expression:

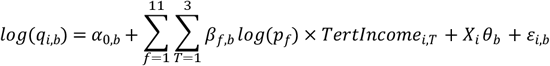

### Baseline intake of beverages

Baseline intake of beverages and self-reported anthropometric data were collected using the POF 2017-2018. Food consumption was assessed using a single 24h recall because two-day information was collected only in a small subsample of participants. We selected individuals 20 years of age and older (37,689 adults), representing 147,852,423 individuals in the adult population in Brazil. Each subject reported the total number of beverages they consumed in standard servings, then we transformed it to milliliters. We selected and classified beverages into SSB, alcoholic beverages, other beverages, and light/diet beverages, according to Table A in S1 Appendix. We added all beverages for each individual in the sample to generate a total beverage consumption variable.

### Reduction in energy intake

To estimate the expected impact of the tax on calories, we created two scenarios considering: 1) the own-price elasticity for SSB by income level, and 2) both own and cross-price elasticities for alcoholic beverages, light diet beverages, and other beverages by income level. We assumed that changes in household purchases derived from price elasticities translated directly into household consumption changes. Prior studies suggest the effects of soda taxes on consumption are linear; thus, we assumed linearity in the changes in consumption for the 20 and 30% tax scenarios.[18]

### Adjustment of self-reporting bias on weight and height

We adjusted self-reported height and weight from POF 2017-2018, linked to SSB consumption data, using measured data from the Brazilian National Health Survey (*Pesquisa Nacional de Saúde* – PNS 2019). The PNS 2019 is a population-based survey, nationally representative, aiming at estimating living and health conditions. This survey measured weight and height using scales, portable stadiometers, and anthropometric tapes with the appropriate training and supervisors. More information about the PNS 2019 can be found elsewhere [19]. Being both POF and PNS nationally representative surveys, we could adjust for self-reporting bias. We adjusted self-reported weight and height from POF to match the distribution of weight and height measured in PNS 2019, following previously published methods.[20] Briefly, we calculated the difference between weight and height data quantiles from both surveys and fitted a cubic spline to smoothly construct a distribution of bias for weight and height. Then, using the quantiles of self-reported weight and height, we estimated the bias using fitted cubic splines (Figures A-B in S1 Appendix). Finally, we added the predicted difference to the self-reported data by sex, as previous articles reported a larger underestimation of weight and overestimation of height in women compared to men.[21] Details are available in section 2 from S1 Appendix.

### Reduction in body weight, BMI, and obesity prevalence

To simulate weight changes attributable to the SSB tax on the Brazilian population, we used a microsimulation approach, a dynamic model developed by Hall et al.[22] This model has been previously implemented to estimate the impact of the sugar-sweetened beverages tax.[9,23,24] The model considers changes in extracellular fluid, glycogen, and fat and lean tissues caused by the change in caloric consumption but keeping the physical activity constant at the individual level. Bodyweight is the result of the sum of fat mass and fat-free mass, and it is determined by the model using a system of ordinary differential equations. A detailed description of the equations and model implementation can be found elsewhere.[22] Dietary and anthropometric information from POF were used as inputs for the model to obtain weight changes over ten years, then translated into BMI changes and obesity prevalence changes. An overview of the simulation process is shown in section 3 from S1 Appendix.

#### Sensitivity analysis

We relied on self-reported weight and height to generate the expected impact of the SSB tax, adjusting the self-report bias expected in POF 2017/2018 using quantiles of weight and height data from PNS 2019. However, recent concerns have been raised about the potential limitations of this method.[25] To validate the results of our analysis, we followed an independent estimation procedure, using measured weight and height from PNS 2019 and SSB consumption from POF 2017/2018. Since these two surveys cannot be linked at the individual level, we estimated the average SSB consumption and BMI for 14 groups by age (20-39, 40-59, 60+) and socioeconomic status (low, middle, and high) for males and females. Then, we implemented Hall’s dynamic model using the averages, generating the expected weight reduction in kilograms for each group and compared against the estimated produced using the corrected self-reported data. Results and methods for this analysis are fully available in section 6 in S1 Appendix.

### Reduction in cases with obesity

We translated the reductions in the prevalence of obesity into obesity cases averted over 10 years using the baseline prevalence of obesity in Brazil (assuming a steady state) and the expected reductions in obesity, multiplied by the population projections from the Brazilian Institute of Geography and Statistics for the adult population from 2021 to 2030.

### Reduction in healthcare costs

Obesity costs were obtained from a 2020 publication with direct costs attributable to obesity in Brazil: US$ 365.7 million in 2018 (1 US$ = R$ 3,875). [26] Direct costs use the health system perspective, in which out-of-pocket expenses are not included. In 2018, we estimated a 25.2% obesity prevalence in the adult population in Brazil using POF, resulting in 37.2 million persons with obesity. Dividing the annual cost of obesity (US$ 365.7 million) by the total number of obesity cases, we estimated an annual cost per capita of US$ 9.84. Using the Consumer Price Index Inflation rate, we converted costs from 2018 to costs in 2021 and obtained an annual cost of US$ 10.34. Finally, we multiplied the absolute yearly reduction in obesity prevalence by the obesity costs to estimate the total direct costs. A detailed description of cost estimations is included in section 5 from S1 Appendix, following the Consolidated Health Economic Evaluation Reporting Standards (CHEERS) (S2 Appendix).

## RESULTS

Table 1 presents the estimated price elasticities for SSBs by income level. Low-income households were, on average, more sensitive to changes in SSB prices, as they had higher price elasticities than higher-income households. SSB price elasticities ranged from -1.13 (high income) to -1.24 (low income). Substitution effects from increases in SSB prices were only observed for other beverages (which included milk, water, and 100% fruit juices) in low and high-income households and for light/diet beverages for high-income households. We observed a complementary behavior between the acquisition of SSBs and alcoholic beverages (if one product decreased, the other also decreased).

**Table 1:** Price elasticities of SSB by income tertiles, 2018.

|  |  | Sugar<br>sweetened<br>beverages | Other<br>beverages | Alcohol<br>beverages | Light/diet<br>beverages |
| --- | --- | --- | --- | --- | --- |
| Dependent variable: Log of Quantity Acquired (in Liters) |  |  |  |  |  |
| Low<br>income | *Mean* | -1.241*** | 0.046*** | -0.122** | -0.174 |
|  | *sd* | -0.015 | -0.017 | -0.053 | -0.124 |
| Middle<br>income | *Mean* | -1.186*** | 0.03 | -0.076 | 0.094 |
|  | *sd* | -0.019 | -0.021 | -0.047 | -0.11 |
| High<br>income | *Mean* | -1.126*** | 0.066*** | -0.141*** | 0.201** |
|  | *sd* | -0.022 | -0.025 | -0.048 | -0.084 |
| *Observations (n)* |  | 18,876 | 27,488 | 4,263 | 383 |

Notes: All regressions used microdata from POF 2017-2018. Income levels were divided using the tertiles of the distribution of the income per capita of the households). Other controls include logarithm of prices of other products (11 categories included), of income (in levels and squared), characteristics of household head (schooling, age, gender, race), total number of household members and total number of children living within the household (all ages). All regressions are weighted using sample survey weights. Standard errors (in brackets) are clustered by socioeconomic strata. P-values: \* 0.10 \*\* 0.05 \*\*\* 0.01

Table 2 presents the baseline intake of SSBs and other beverages among Brazilian adults in 2018. We estimated a baseline intake of 72 kcal/person/day from SSB. The expected caloric change after the SSBs tax was estimated to be -16.9 kcal/person/day for 20% and -25.3% kcal/day for 30% tax. If we included changes in other beverages to allow for substitution, expected caloric changes increased to -17.3 kcal/person/day for 20% (an extra reduction of 0.4 kcal) and -25.9 kcal/person/day for 30% tax (an extra reduction of 0.6 kcal). Expected caloric changes should be higher among younger than older adults and higher income levels.

**Table 2.** Baseline caloric intake from SSBs and other beverages (alcohol, light and other beverages)

|  | Populati<br>on size<br>(million) | Baseli<br>ne<br>intake<br>kcal/<br>day | 95% CI | Caloric changes from SSB |  |  |  | Other caloric changes |  |  |  |
| --- | --- | --- | --- | --- | --- | --- | --- | --- | --- | --- | --- |
|  |  |  |  | 20% tax |  | 30% tax |  | 20% tax |  | 30% tax |  |
|  |  |  |  | Mean<br>change<br>Kcal/<br>day | 95% CI | Mean<br>change<br>Kcal/<br>day | 95% CI | Mean<br>change<br>Kcal/<br>day | 95% CI | Mean<br>change<br>Kcal/<br>day | 95% CI |
| Total | 147.9 | 71.8 | 69.3,74.4 | -16.9 | -17.5,-16.3 | -25.3 | -26.2,-24.4 | -0.4 | -0.4,-0.3 | -0.6 | -0.7,-0.5 |
| sex |  |  |  |  |  |  |  |  |  |  |  |
| *female* | 78.2 | 65.1 | 62.4,67.7 | -15.3 | -15.9,-14.7 | -22.9 | -23.9,-22.0 | -0.1 | -0.1,-0.0 | -0.1 | -0.2,-0.0 |
| *male* | 69.6 | 79.5 | 75.8,83.1 | -18.6 | -19.5,-17.8 | -27.9 | -29.2,-26.7 | -0.7 | -0.8,-0.6 | -1.0 | -1.2,-0.9 |
| Age groups |  |  |  |  |  |  |  |  |  |  |  |
| 20-39 | 62.2 | 92.2 | 88.0,96.5 | -21.7 | -22.7,-20.7 | -32.6 | -34.1,-31.1 | -0.4 | -0.5,-0.3 | -0.7 | -0.8,-0.5 |
| 40-59 | 53.9 | 62.9 | 59.4,66.4 | -14.7 | -15.5,-13.9 | -22.1 | -23.3,-20.8 | -0.5 | -0.6,-0.3 | -0.7 | -0.8,-0.5 |
| 60+ | 31.7 | 46.9 | 43.3,50.6 | -11.0 | -11.9,-10.2 | -16.6 | -17.8,-15.3 | -0.1 | -0.2,-0.0 | -0.2 | -0.3,-0.1 |
| Income status |  |  |  |  |  |  |  |  |  |  |  |
| Low | 47.4 | 53.5 | 49.9,57.1 | -13.3 | -14.2,-12.4 | -19.9 | -21.3,-18.6 | -0.2 | -0.3,-0.2 | -0.4 | -0.5,-0.2 |
| Middle | 49.1 | 74.0 | 70.2,77.8 | -17.5 | -18.5,-16.6 | -26.3 | -27.7,-25.0 | -0.2 | -0.3,-0.2 | -0.3 | -0.4,-0.2 |
| High | 51.4 | 86.7 | 81.7,91.8 | -19.5 | -20.7,-18.4 | -29.3 | -31.0,-27.6 | -0.6 | -0.8,-0.5 | -0.9 | -1.2,-0.7 |

Table 3 presents the expected impact of the SSB tax over the prevalence of obesity, 10 years after implementation, using own-price elasticities. The obesity prevalence is expected to decrease from 25.2% to 23.6%, resulting in a 6.3% reduction with a 20% tax and an 8.7% obesity reduction with a 30% tax. This reduction is expected to be higher among younger (−8.3%) than older adults (−3.6%.) and among higher (−7.8%) than lower-income levels (−5.4%). Allowing for substitution using cross-price elasticities, the expected decrease in the obesity prevalence becomes larger, being -6.7% for 20% and -9.1% for a 30% tax.

**Table 3.**
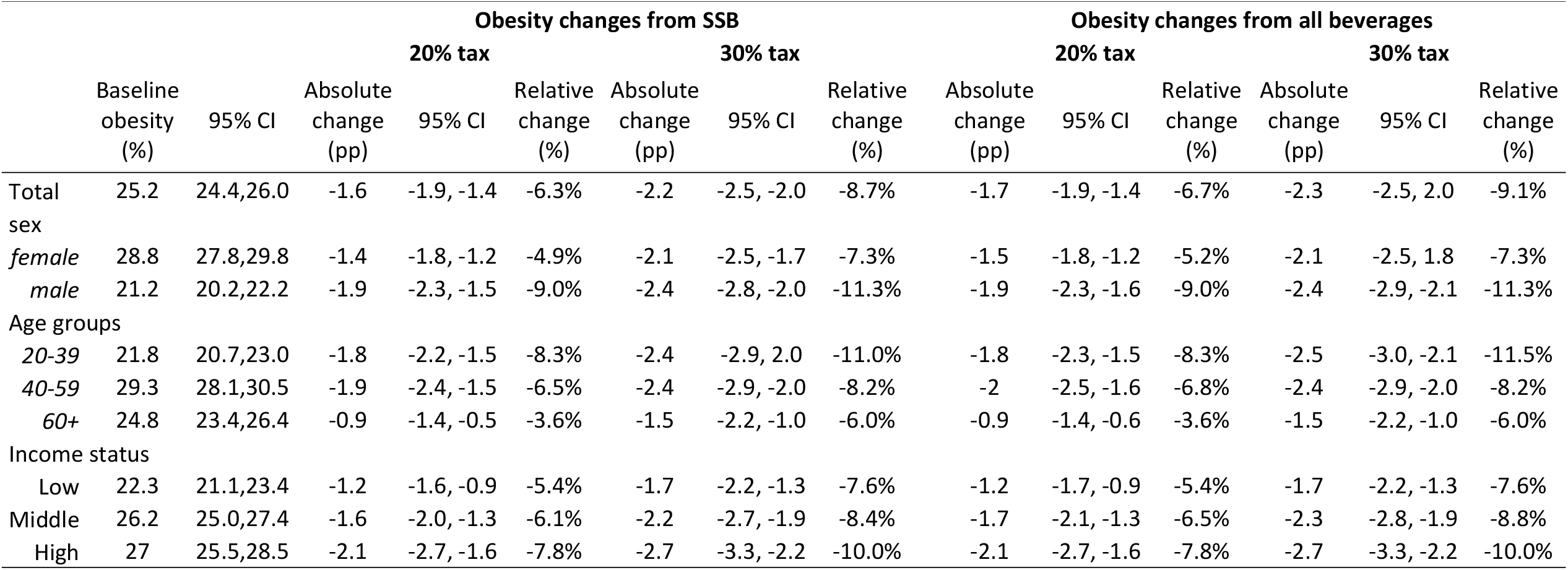
Baseline obesity in Brazil and expected obesity reduction after SSB tax, considering caloric changes from SSBs and all caloric changes.

Fig 2 shows the expected obesity reduction in cases and potential cost savings 10 years after implementing the SSB tax, using own-price elasticities. A 20% tax is expected to reduce the number of people living with obesity by 2.8 million and by 3.7 million with a 30% tax; this would translate into direct cost savings of $US 253.9 million with a 20% tax and $US 341.8 million with a 30% tax. Using cross-price elasticities, these estimates increase, so that the reduction in the number of people living in obesity reaches -2.8 million and -3.8 million for a 30% tax; cost savings increase to $US 257.4 million with a 20%, and $US 345.9 million with a 30% tax.

**Fig 2.**
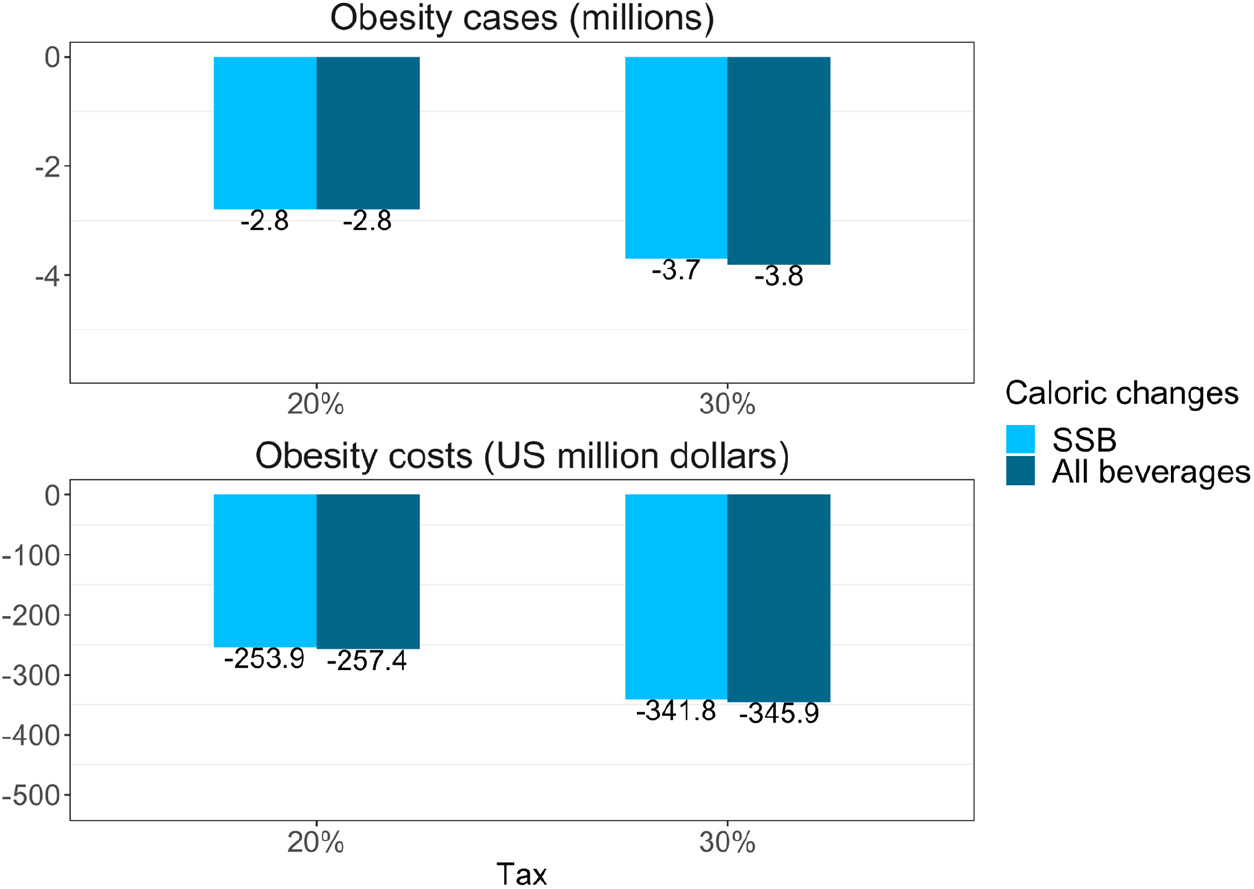
Reduction in cases and cost savings 10 years after implementing the SSB tax, using own-price elasticities if we consider only the effect on SSBs or all beverages.

## DISCUSSION

We aimed to estimate the own and cross-price elasticities of SSBs and translate them into the expected impact of a 20 and 30% SSB tax on obesity over 10 years in Brazil. In Brazil, we found that SSB is elastic and weakly substituted by milk, water, 100% fruit juices, and light beverages. Low-income households were more sensitive to price increases and had a higher elasticity than other income levels. We estimated that a 20% tax on SSBs could translate into a 6.7% reduction in the obesity prevalence and US$ 257.4 million savings in obesity costs over ten years after implementation. A 30% tax was estimated to reduce 9.1% the obesity prevalence and US$ 345.9 million in the obesity costs over ten years. The largest benefits of the tax are expected to be experienced by younger adults and by people in the high-income tertile.

Estimating the impact of price increases on demand for SSB and other beverages in Brazil is key to assessing the potential impact of an SSB tax across income strata. Our analysis indicates that a 10% price increase to SSB in Brazil should result in a 12.4% reduction in purchases among low-income households, 11.9% in the middle, and 11.3% in the high-income strata; this is similar to what has been previously estimated in Mexico, where a 10% price increase was expected to produce an 11.6% reduction in purchases, but higher than a previous analysis in Brazil, that estimated that a 10% price increase would result in an 8.5% reduction in SSB, using data from 2002-2003.[27,28] Our estimates are also similar to the findings of a meta-analysis of observational studies that evaluated changes in intake and consumption after a 10% tax that showed an associated 10% reduction in SSB purchases and intake.[5] Our analysis suggests that SSB purchases in Brazil will decrease if prices are increased, opening the possibility for an SSB tax to produce an impact on consumption.

We also analyzed cross-price elasticities, finding only a weak substitution by milk, water, and 100% fruit juices. Substitution for these beverages was previously reported in a meta-analysis of US, Mexico, France, and Brazil studies.[29] In contrast, our findings suggest that alcohol purchases are expected to decrease if SSB prices increase, while non-caloric beverages will increase only in high-income adults. Considering these substitutions, a 20% tax should lead to a direct caloric reduction of 16.9 kcal from SSB, which will be further increased by substitution or complementation for other liquids with a lower caloric density (−0.4 kcal after a 20% tax and -0.6 kcal after a 30% tax). This finding implies that if an SSB tax were to be implemented, no caloric substitution for liquids is to be expected, an observation that is in line with a prior observational study that found a weak caloric compensation for SSB in the Brazilian population[30] and with international studies that suggest that caloric compensation for liquids is incomplete.[31]

This is the first study in Brazil to estimate the potential impact of a 20 and 30% SSB tax on body weight. Based on our modeling exercise, we estimated that a 20% tax could result in a 6.7% reduction in obesity, increasing to 9.1% under a 30% tax. This estimate is within the range of prior modeling studies using a comparable approach. For instance, in Mexico, a 20% tax was estimated to reduce 6.9% the obesity prevalence after ten years.[9] We observed important differences in the expected obesity impact by income tertiles; high-income groups are expected to reduce their obesity prevalence by 7.8%, compared to 5.4% in low-income groups. This could be explained by differences in baseline consumption of SSB, which were 86.7 kcal/day for high-, 74.0 kcal/day for middle-, and 53.5 kcal/day for low-income groups. Also, baseline obesity prevalence was 27.0% in high, 26.2% in the middle, and 22.3% in low-income groups; which, along with greater baseline SSB consumption, explain the greater expected reductions in BMI among high-income individuals. We found that a 20 and 30% excise tax on SSB will produce benefits for all income groups as it will reduce obesity levels in all SES groups, however an augmented tax revenue can more disproportionately benefit lower-income groups if they are used to fund redistributive social policies, the Brazilian national unified health system, in particular strategies associated with obesity prevention and treatment.[32]

An SSB tax in Brazil is expected to produce important savings related to obesity prevention. We estimated a reduction of US$ 257.4 million in obesity costs ten years after the implementation of a 20% SSB tax. This cost reduction is smaller than what has been estimated in the US (US$ 23.6 billion) but higher than for Mexico (US$ 159.5 million) for a similar tax increase (20%) and a similar period (ten years).[12,33] Cost savings estimated in our analysis do not consider out-of-pocket and indirect costs of obesity, such as those related to loss of productivity, absenteeism, or disability. Indirect costs for diabetes were estimated to be 70% of the total diabetes costs in Brazil,[34] which suggest that our estimates of obesity-related cost savings are conservative. Finally, our cost-analysis does not consider the potential benefits derived from the social investment of the tax revenue, particularly if revenue is directed at strengthening obesity prevention measures, which can further increase the health benefits from the tax.

Some limitations of our study should be mentioned. Available baseline weight and height were self-reported. We relied on a quantile distribution method to calibrate it to the PNS objectively measured data (section 2, S1 Appendix); yet, this method is imperfect and could be insufficient to correct reporting bias.[25] For that reason, we performed a sensitivity analysis using the same dynamic model, but at aggregated level. In this analysis, we used objectively measured weight and height from 14 sex, age, and SES groups (aggregated data) and linked it to SSB consumption. Differences in the expected weight reduction associated with the SSB tax were minimal. In the 14 groups, the aggreged estimation fell within the confidence intervals for the individual model (Table H in S1 Appendix) This sensitivity analysis increases our confidence in the self-reported correction to estimate weight reductions in kilograms, but a similar approach could not be used to assess the effect of the correction on the BMI classification. However, we estimated a 16.9% obesity prevalence using self-reported weight and height, compared to 25.2% using adjusted self-reported data, and to 26.8% obesity prevalence using objective measures of weight and height (Table B in S1 Appendix). The potential error of the adjusted self-reported data on the BMI classification (obesity vs non-obesity) in comparison with the measured data is in the downward direction, suggesting that adjusted data could underestimate the SSB impact on obesity, but in a lower magnitude than if we had used self-reported data without adjustments.

Because the available literature in obesity costs in Brazil are very heterogeneous, our findings on cost reduction also have limitations. A systematic review found that obesity costs in Brazil (including direct and indirect costs) ranged from USD 133.8 million to USD 6.3 billion per year (47 times the lowest estimation).[35] We relied on the most recent obesity cost estimation in Brazil that uses attributable fraction formulas to calculate the attributable costs to obesity, including 26 diseases (USD 365.7 million per year), and lies between the cost range of the systematic review.[26] Still, the cost estimation uses a health-system perspective, so out-of-pocket and indirect costs are not included, leading to a conservative estimate of the economic benefits of the tax. Our model relies on a steady-state assumption, which implies that the obesity prevalence is constant over the 10 years, and the only caloric change observed is the caloric reduction produced by the SSB tax. If obesity continues to rise, failing to include an increasing trend would lead to an underestimation of the total number of obesity cases reduced and obesity costs because the estimated reduction over time would be multiplied by a fewer number of obesity cases.

Sugar-sweetened beverage taxes have been implemented in more than 73 countries worldwide.[7] Studies have shown that SSB are elastic and respond to increases in price; yet no negative economic effects have been observed in countries where SSB taxes have been implemented,[32] and analyses in Brazil suggest an economic net benefit from implementing an SSB tax.[36]Our findings suggest that implementing an SSB tax in Brazil will lead to an important reduction in the caloric intake of beverages with little to no nutritional added value; this reduction could help reduce the obesity prevalence across all socioeconomic strata. From an economic perspective, an SSB tax could reduce healthcare costs while generating revenue that could be in turn invested in other obesity prevention interventions. In 2017, the World Health Organization recommended that all countries should implement an SSB tax of at least 20%. Our analysis suggests that a 30% tax in Brazil could produce greater population health benefits for all income strata.

## Data Availability

All relevant data are within the manuscript and its Supporting Information files.

## Notes

DISCLOSURE: The authors declared no conflict of interest.

### Competing Interest Statement

The authors have declared no competing interest.

### Funding Statement

ABA,RTA, TBG, PP, ACD are funded by a grant awarded by Bloomberg Philanthropies through a sub award agreement (5104695) between the University of North Carolina at Chapel Hill and the University of Sao Paulo Center for Epidemiological Studies in Nutrition and Health, Brazil. The funder had no role in study design, data collection and analysis, decision to publish, or preparation of the manuscript.

